# HARMONY (HARM reduction for Opiates, Nicotine and You): Statistical Analysis Plan for a Randomised Controlled Trial of the Effectiveness of Vaporised Nicotine Products for Tobacco Smoking Cessation amongst NSW Opiate Agonist Treatment Clients

**DOI:** 10.1101/2024.06.21.24309282

**Authors:** C Oldmeadow, E Nolan, B Bonevski, M Jackson, N Lintzeris, N Ezard, C Gartner, P Haber, R Hallinan, C Rodgers, T Ho, A Dunlop

## Abstract

**Background:** The HARMONY study is a multicentre, randomised, single-blinded parallel group trial. It will compare the effectiveness of a 12-week course of liquid nicotine delivered via vapourised nicotine products (VNPs) to best practice nicotine replacement therapy (NRT) for smoking cessation in individuals receiving opiate agonist treatment (OAT).

The aim of publishing this statistical analysis plan is to make the pre-specified statistical principles and procedures to be performed in the analysis of data generated by the HARMONY study, publicly accessible prior to the commencement of data analysis.

**Methods:** The plan outlines the analysis procedures for analysing the primary outcome of self-reported 7-day point prevalence abstinence from tobacco after 12-weeks of treatment. Secondary outcomes include biochemically verified abstinence, self-reported 30-day abstinence, number of cigarettes smoked each day, craving and withdrawal symptoms, and VNP safety. Between-group comparisons will be conducted at end of treatment, and at 12-weeks post-treatment. Researchers collecting outcome data are blind to the treatment group of each participant.

**Analysis:** Bayesian hierarchical models will be used to estimate the treatment effects for all outcomes with uninformative prior distributions for all effect parameters. Alongside the treatment effect estimate of each outcome, a 95% credible interval (highest posterior density), Bayes factor, and probability of direction will be presented. The analyses will be performed under an ITT framework assuming missing at random. All missing outcome and baseline data will be multiply imputed with predictive mean matching.

**Conclusion:** Making the statistical analysis plan for the HARMONY study publicly accessible prior to the commencement of data analysis minimises the risk of bias in the analysis of data, and the interpretation and reporting of results generated by the study.

**Registration:** The study was registered in the Australian New Zealand Clinical Trials Registry (Reference ACTRN12621000148875).

## INTRODUCTION AND OBJECTIVES

### Background and rationale

- Smoking continues to be the main cause of preventable disease in Australia linked to cancer, respiratory disease and cardiovascular disease. Although smoking rates have decreased in the general population in Australia (13%), they continue to be disproportionately high amongst lower socioeconomic status groups.
- Tobacco smoking is highly prevalent amongst opiate agonist treatment clients. The highest rates of smoking amongst people with substance use disorders are seen in those on opiate agonist treatment (OAT): as high as 73% to 94% smoking prevalence.
- Clients in OAT are greatly interested in quitting smoking, especially if provided in their treatment clinic.
- While smoking cessation appears possible in OAT smokers, few interventions have demonstrated efficacy in this population of tobacco users.
- Most NRT interventions decreased cigarette use, but sustained abstinence rates remained low. High relapse rates in this population may be due to factors related to addiction (e.g. nicotine enhancing the opioid withdrawal reductions due to OAT), lack of cessation support, and the high levels of smoking in their social network.
- Vaporised nicotine products (VNPs) are a broad range of battery-powered devices that deliver an aerosol of propylene glycol and/or glycerine, nicotine and flavours.
- Large scale population surveys in the US (n=161,054) and UK (n= 170,490) suggest that VNP use is significantly associated with smoking cessation.
- A small (n=12), single group trial in the USA among people on OAT found reductions in cigarettes smoked per day over a 6 week, VNP-use study period. There are no other trials of VNPs for smoking cessation amongst OAT clients published or registered.

### Objectives

#### Primary objective

1. The primary aim of the study is to examine the effectiveness of treatment with VNP compared to best practice NRT on self-reported 7-day point prevalence from tobacco smoking at the end of 12 weeks of nicotine treatment in OAT clients.

#### Secondary Objectives

1. To compare the effectiveness of treatment with VNP compared to best practice NRT on biochemically verified self-reported 7-day point prevalence abstinence from tobacco smoking at the end of 12 weeks of nicotine treatment in OAT clients.
2. To compare self-reported 30-day continuous abstinence from tobacco smoking for the VNP treatment group relative to the best practice NRT group at 3 months post-treatment.
3. To compare self-reported 7-day point prevalence abstinence from tobacco smoking for the VNP treatment group relative to the best practice NRT group at 3 months post-treatment.
4. To examine the safety of liquid nicotine delivered via VNPs in the study population, relative to best practice NRT.
5. To determine if treatment with VNP relative to best practice NRT reduces the number of cigarettes smoked per day by OAT clients prior to treatment by 50% (or more) at end of treatment and 3 months post-treatment.
6. To compare nicotine craving and withdrawal symptoms for participants in the two treatment groups (VNP vs best practice NRT) at end of treatment and 3 months post-treatment.
7. To compare tobacco relapse episodes for participants in the two treatment groups (VNP vs best practice NRT) at end of treatment and 3 months post-treatment.
8. To compare nicotine treatment adherence for participants in the two treatment groups (VNP vs best practice NRT) at end of treatment and 3 months post-treatment.
9. Compare other drug use in the two groups (including cannabis use, amphetamine, nonprescribed opioids) at end of treatment and 3 months post-treatment.
10. To compare study retention for participants in the two treatment groups (VNP vs best practice NRT) at end of treatment and 3 months post-treatment.
11. A within trial analysis to compare the cost and consequence of treatments for participants in the two arms (VNP vs best practice NRT).

### Hypotheses

The primary hypothesis is that treatment with VNP has a greater 7-day point prevalence of abstinence to tobacco smoking than best practice NRT. Where appropriate, the study will compare outcomes under a superiority hypothesis, however, it may be appropriate to consider non-inferiority hypothesis when considering the outcomes related to safety, cravings and adherence.

## METHODS

### Trial design

A two-arm randomised single blind trial comparing a 12-week course of liquid nicotine delivery via VNP’s compared to best practice NRT. The two treatments are randomised in a 1:1 ratio of treatment arm allocation.

#### Condition 1: VNP

Participants randomised to Condition 1 shall be supplied with a VNP (Innokin Endura T18-II starter kit) and 12 weeks of prescribed liquid nicotine (Nicophar). A one-week supply of NRT transdermal patches shall also be provided for use whilst they are learning how to use the VNP effectively.

#### Condition 2: NRT

Participants randomised to Condition 2 shall be supplied with a combination of NRT transdermal patches and oral forms (inhalators, gum, lozenges, mouth spray) to use throughout the 12-week intervention period.

### Randomisation

The randomisation schedule was developed by an independent statistician and conducted in REDCap. The randomisation was stratified, through blocking, by treatment site and cannabis use.

### Sample size

A sample of 200 smokers in each treatment group were needed to detect a difference of 6% between groups (i.e., 1% in NRT group and 7% in VNP group continuous abstinence at 6-month follow-up) with 80% power and a 5% significance level. Based on our previous research in AOD setting with clients from various programs including OAT and assuming 30% attrition rate at 6 months follow-up, we required a sample size of 572 eligible smokers across 6 OAT sites

### Framework

This is a superiority trial.

### Statistical interim analyses and stopping guidance

No interim analyses are planned.

### Timing of final analysis

The primary analysis will occur at the completion of the trial, once all participants have completed the three-month (24 – 28 weeks) follow-up or are determined to be lost to follow-up.

### Timing of outcome assessments

Outcome data will be collected from participants at two time-points: (i) at baseline following consent; (ii) at the end of the 12-week treatment and, (iii) 3-months post treatment over the phone by a member of an independent CATI team. See Figure 1 for participant flow through the study.

**Figure 1.**
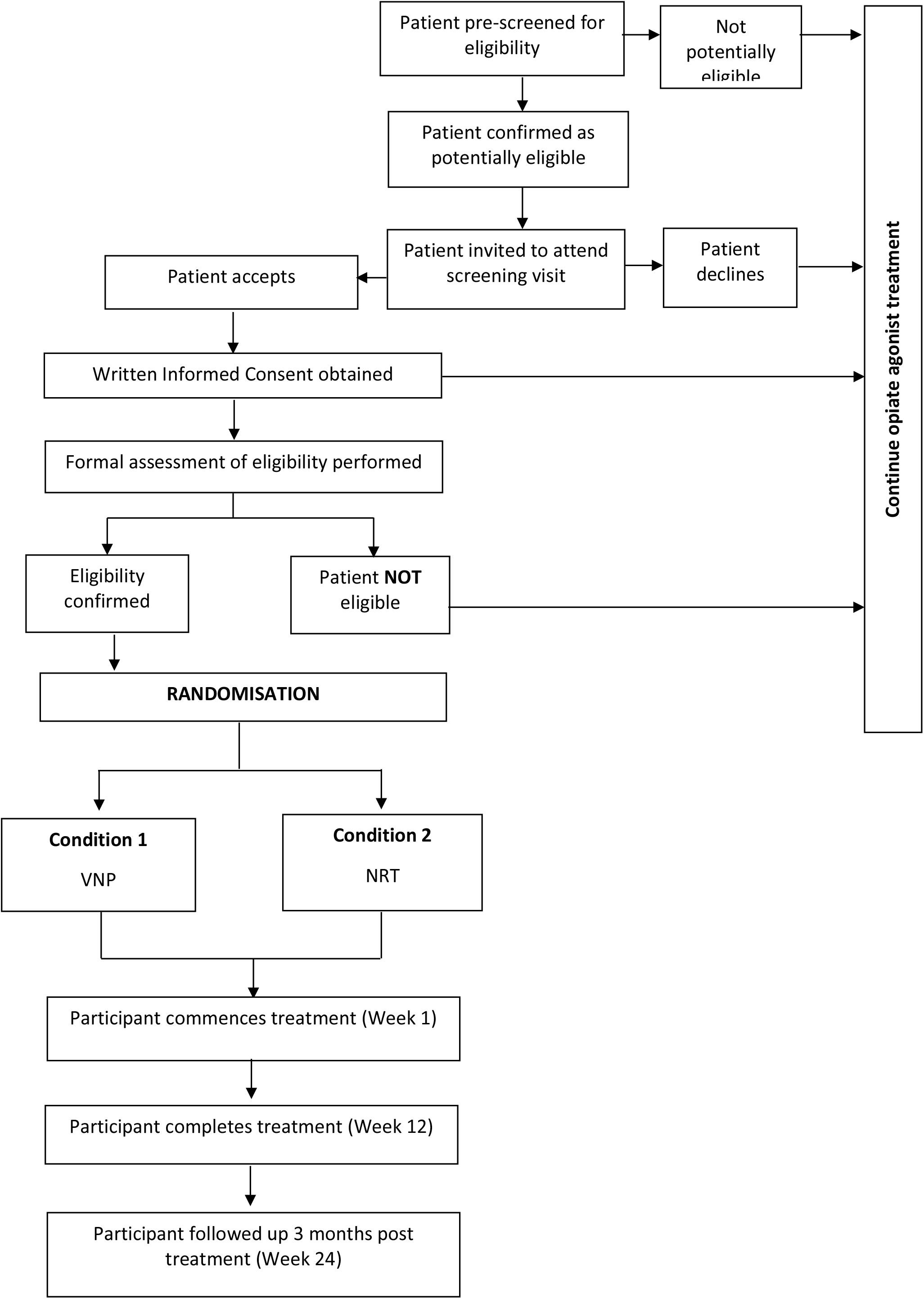
**CONSORT diagram of participant flow through the study.**

## TRIAL POPULATION

### Screening data

A RO/RN will complete a pre-screening questionnaire with the client either in person or by telephone. Potential eligible persons will be invited to attend the clinic in person or via Telehealth for an informed consent discussion and a formal screening assessment.

### Eligibility

Patients will be eligible to participate if the following apply:

- Provide written, informed consent to participate in the study.
- Aged 18 years or older
- Be accessing opioid agonist treatment from a participating service
- Current daily tobacco smokers on self-report
- Want to quit or cut down their smoking
- Be willing and able to comply with requirements of study (including having access to a phone)

### 1.1. Exclusion Criteria

Patients will be ineligible to take part in the study if the following apply:

- Currently breastfeeding or pregnant, or of childbearing potential and planning/trying to fall pregnant during the study period.
- Current, severe medical disorder assessed by study medical officer (such as but not limited to, unstable cardiovascular/peripheral vascular disease, poorly controlled hypertension).
- Current, severe or unstable psychiatric disorder assessed by study medical officer (such as but not limited to, acute psychosis, severe anxiety and/or mood disorder, intent to harm self or others).
- Current enrolment in a clinical trial involving any investigational drug.
- Regular use (more than one day per week) of Vaporised Nicotine Product (or e-Cigarette) containing nicotine in the last 30 days.
- Not available for follow-up (e.g. likely imprisonment or transfer out of service to another service that is not a trial recruitment site).

### Recruitment

The recruitment strategy for this study will incorporate approved advertisements being displayed in participating services OAT waiting rooms and other suitable locations. Due to recent changes in OAT service delivery, advertisements may also be sent via text or email to clinic clients that are no longer visiting clinics daily. Interested clients may contact the researchers directly using contact details contained in the advertisements.

Introductions to the trial shall also be made by the clinicians at each site during appointments or conversations. Clinicians will be asked to identify clients who smoke and enquire about their desire to stop and their interest in participating in a smoking cessation treatment using the script provided. The contact details of any interested clients will be provided to the research team for pre-screening.

### Withdrawal/follow-up

Treatment discontinuation does not equate withdrawal from the study and these participants will be invited to continue completing the research component of the study.

### Involuntary discontinuation

Participants may be withdrawn involuntarily by the investigator (or delegate) if they meet the following criteria:

- Participant experiences a severe or serious adverse event, thought to be related to the study drug/device, which is not resolving.
- Absence from the protocol monthly visits (Week 4, 8 or 12) for more than four weeks.

Even if the participant has capacity to consent at baseline, the intervention staff will assess capacity to consent at each contact and will cease the intervention if the participant is deemed to lack capacity. In this case the reasoning will be explained and recorded on the CRF.

### Voluntary discontinuation

Participants have the option to stop treatment or revoke their consent at any time without giving a reason. The distinction between stopping treatment and revoking consent is shown by the following definitions:

- Discontinuation of treatment: A participant would be considered to have discontinued protocol treatment if they stop using the VNP device/NRT medication study. In this case the participant will continue in all remaining research interviews and assessments.
- Revocation of consent: Total withdrawal from the trial would occur in the circumstance that the participant decides to revoke their consent. Under these circumstances no further information would be collected from the participant for the purpose of the trial.

Participants may at any time elect to revoke consent for study participation without jeopardising their relationship with either their doctor or treatment service.

## STATISTICAL PRINCIPLES

### General Statistical methodology

Statistical inference for assessing differences between groups for all outcome measures will be within a Bayesian framework. This involves for each outcome specifying a likelihood function, and a prior distribution. Together these form the posterior distribution for the parameters of interest.

#### Prior specification

The prior distribution for all treatment effect parameters of interest will be an uninformative prior and all other effect parameters will have an uninformative prior. These are specified in detail for each outcome below.

#### Samples from the posterior distribution

Samples from the posterior distribution will be obtained using the No U-Turn Sampler (NUTS) [1], as implemented in the brms R package [2]. All models will be run with 2,000 burn-ins and 10,000 iterations (across 4 chains). A thinning rate of 1 will be used.

#### Assessment of convergence

To assess convergence, a visual inspection of chains and histograms of posterior distributions for all model parameters, and Gelman and Rubin’s Rhat will be examined. An Rhat <=1.1 indicates chains have converged. If chains have not converged then we will initially attempt to increase the number of iterations, and then try stronger prior distributions.

#### Assessment of fit

To assess model fit, the posterior predicted values of the outcome will be compared to the observed data.

#### Summaries of the posterior distribution

The mean of the converged posterior distribution for the parameter reflecting the between group differences (either absolute or relative) will be presented as the point estimate to 2 decimal places. 95% credible intervals will be calculated using the highest posterior density (HPD) method and presented to 2 decimal places. The one-sided probability that the difference between treatment and control is greater than zero (representing a beneficial treatment effect) will be estimated from the posterior distribution as the proportion of posterior samples that are of the sign that is favorable to the intervention (either +’ve or –‘ve).

#### Bayes factor

The Bayes factor will be computed using the Savage-Dickey density ratio method [3] in the brms package [2]. The Bayes factor is interpreted as the multiplicative likelihood of the alternative hypothesis compared to the null hypothesis after considering the data. A value greater than one indicates more support for the alternative hypothesis compared to the null.

### Analysis populations

The primary analysis modelling will be conducted as intention-to-treat (ITT). The ITT sample will include all participants who are randomised (excluding those who choose to withdraw data from analysis), with their data analysed according to the experimental group to which they were randomised to. Descriptive analysis of study data will be performed for all participants who complete at least one time point.

## ANALYSIS

### Baseline patient characteristics

Baseline characteristics will be summarized descriptively using means, standard deviations, medians and interquartile ranges for continuous data, and frequencies and percentages for categorical data. The following baseline characteristics will be presented by treatment arm:

**Table 1.**
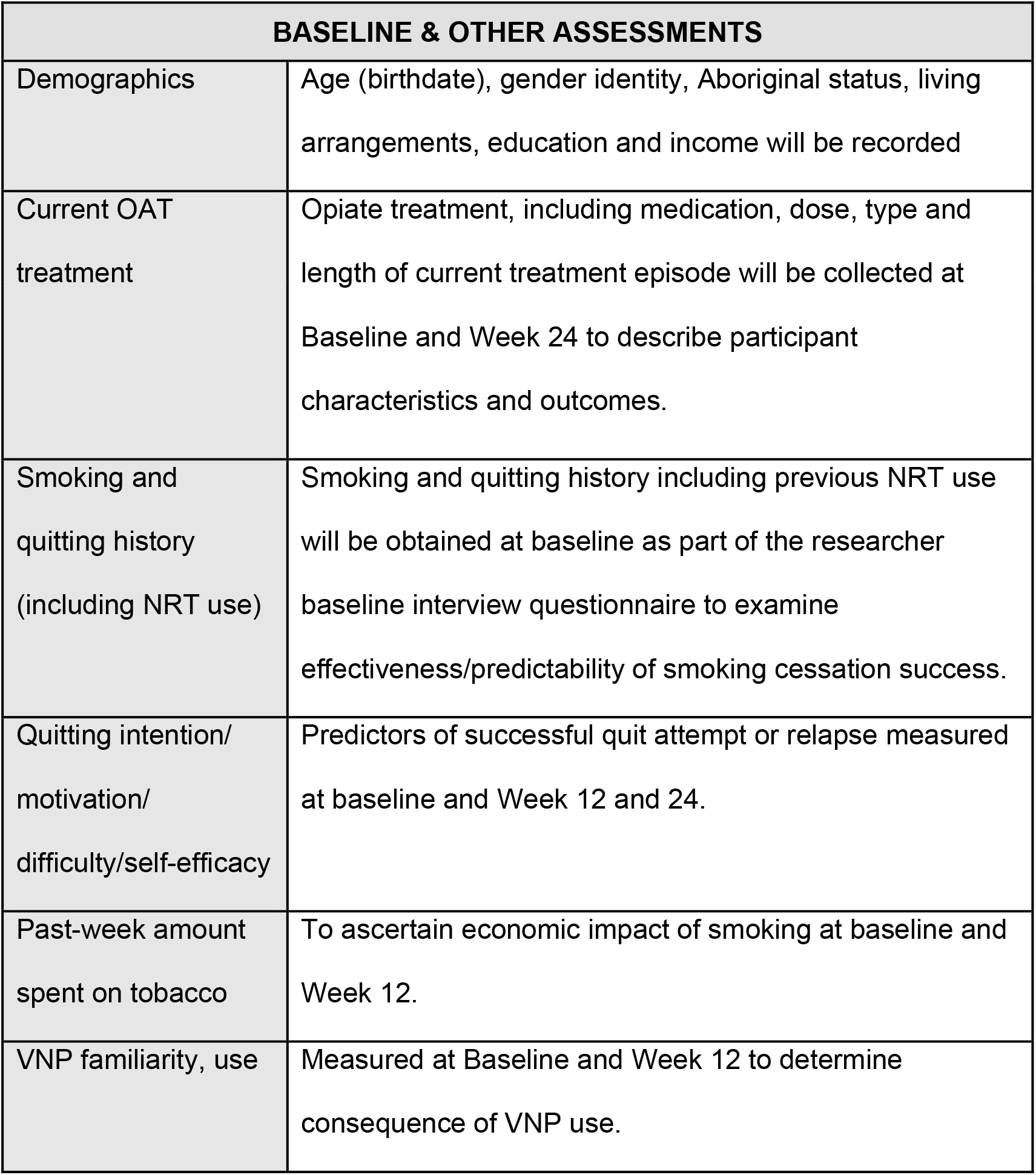
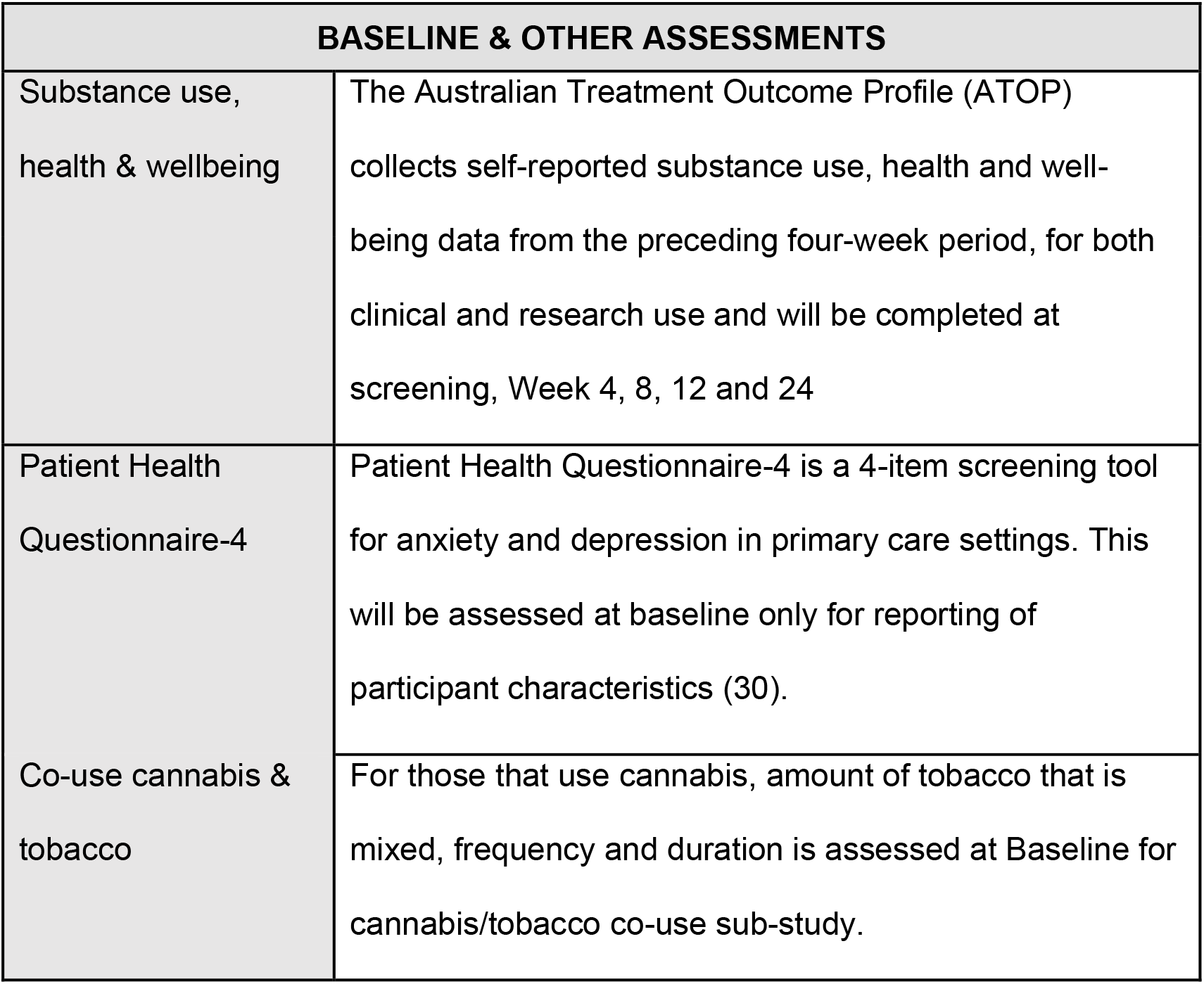
Description of baseline characteristics.

### Outcome definitions

All primary, secondary outcomes will be measured at baseline, at the end of the 12-week treatment and 3-months after treatment. An overview of the outcomes to be assessed are shown in Table 2.

**Table 2.** Description of outcomes to be collected and analysed.

| OUTCOME |  |  | QUESTION |
| --- | --- | --- | --- |
| **Primary outcome** | **1** | Self-report 7-day point prevalence abstinence | 7-day point prevalence abstinence from tobacco smoking will be self-reported and based on the following dichotomous question pilot tested in the QuitNic trial (31), “Have you smoked at all, even a puff, in the last 7 days?” Completed at Week 12 |
| **Secondary Outcomes** | **1** | Carbon monoxide (CO) in expired air | Self-report 7-day point prevalence abstinence at end of treatment will be verified using a measure of carbon monoxide (CO) in expired air. Completed at Week 12. |
|  | **2** | Self-reported 30-day continuous abstinence from tobacco smoking | 30-day continuous abstinence from tobacco smoking will be self-reported and based on the following dichotomous question pilot tested in the QuitNic trial (31) “Have you smoked at all, even a puff since the week 12 research interview?” The quantity of cigarettes smoked in the last 30 days will be assessed amongst those who smoked. Completed Week 24. |
|  | **3** | Self-reported 7-day point prevalence abstinence from all tobacco smoking | Self-reported 7-day point prevalence abstinence from nicotine all (including NRT and VNP) will be measured by asking participants "Have you smoked at all, even a puff, in the last 7 days?" Completed Week 24. |
|  | **4** | Adverse Events | Adverse events shall be assessed and recorded from signing the consent form until Week 12. |
|  | 5 | Heaviness of Smoking Index (HSI) | (i) Time to the first cigarette of the day (5, 6–30, 31–60 and 61 + min) and (ii) Average daily consumption of cigarettes (1–10, 11–20, 21–30, 31 + cigarettes). Both completed at Screening and Baseline, Weeks 4, 8, 12 and 24. |
|  | 6 | Craving strength and frequency | Cravings to smoke will be assessed by one item based on Taggar et al (33) “Currently, how often do you get strong cravings to smoke tobacco?” with the response options of 1) Hourly or more often; 2) Several times per day; 3) At least once a day; and 4) Less than daily. Strength will be assessed by asking “How strong have these urges been?” with six response options ranging from No urge to Extremely Strong. Completed at Baseline, Week 12 and 24. |
|  | 6 | Minnesota Nicotine Withdrawal Scale | Nicotine withdrawal will be assessed by the Minnesota Nicotine Withdrawal Scale (34), an eight item scale rating withdrawal symptoms on an ordinal scale ranging from 0 (not present) to 3 (severe). Completed at Baseline, Week 12 and 24. |
|  | 7 | Quit attempts & relapse | Time to relapse and number of quit attempts will be assessed among those who relapsed. Completed at Baseline, Week 4, 8, 12 & 24. |
|  | 8 | Nicotine product adherence | Current use of short and long-acting NRT and e-cigarettes is assessed at Week 4, 8, 12 & 24. At weeks 12 & 24, additional questions around plans for long-term use, length of use and reasons for stopping use are also asked. |
|  | 9 | Substance use, health & wellbeing | The Australian Treatment Outcome Profile (ATOP) collects self-reported substance use, health and well-being data from the preceding four-week period, for both clinical and research use and will be completed at screening, Week 4, 8, 12 and 24. |
|  | 10 | Retention | Compare study retention for participants in the two treatment groups (VNP vs best practice NRT) at end of treatment and 3 months post-treatment |
|  | 11 | Health Economics<br>Cost Analyses | A cost consequence study setting out a detailed comparative cost of treatments - from the perspective of health care provider and primary and secondary outcomes of the VNP and NRT. Costing items include public sources, wage awards, the Medical Benefits Schedule (MBS) and the Pharmaceutical Benefits Scheme (PBS); and research records such as costs of VNP products, inhalators, nicotine patches and gum, batteries and quitting aid materials. Assessed at Week 12. |

### Analysis methods

#### Primary outcome

A logistic hierarchical model will be used to measure the impact of VNP on the primary outcome, self-reported 7-day point prevalence. The model will include a random intercept (assumed normally distributed on the logit scale, with a constant variance) to account for repeated measures within the same participants, a random intercept for site (assumed normally distributed on the logit scale, with a constant variance), fixed effects for baseline self-reported abstinence, cannabis use, time (12-weeks and 3 month follow up), treatment group, and an interaction term for study time and treatment group. Treatment effects will be reported as estimated odds ratio between the two treatment groups at each timepoint with 95% credible intervals (Highest Posterior Density) and a bayes factor. The results of the primary outcome will be presented in the style of Table A1 (Appendix).

The hierarchical model is specified below:

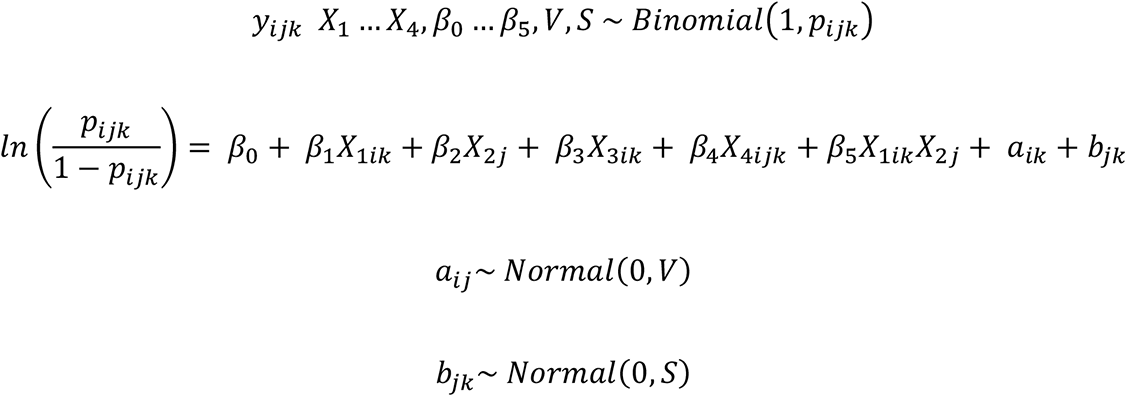

*X_1ik_* = *Treatment group*

*X_2k_* = *Time*

*X_3ik_* = *Baseline self reported abstinence*

*X_4ijk_* = *Cannabis use*

*i* = *participant*

*j* = *time*

*k* = *cluster*

Priors

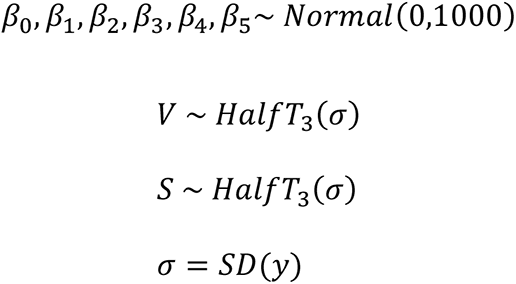

### Secondary outcomes

Hierarchical models with appropriate distributional assumptions will be used to analyse secondary outcomes.

#### Binary outcomes

Logistic hierarchical models with a random intercept (assumed normally distributed on the logit scale, with a constant variance) to account for repeated measures within the same participants, a random intercept for site (assumed normally distributed on the logit scale, with a constant variance), fixed effects for the outcome at baseline, cannabis use, time (12-weeks and 3 month follow up), treatment group, and an interaction term for study time and treatment group. Treatment effects for each follow-up timepoint will be reported as estimated odds ratio between the two treatment groups with 95% credible intervals and a bayes factor.

- (1) & (3) 7-day point prevalence,
- (2) self-reported 30-day continuous abstinence from tobacco smoke,
- (8) adherence,
- (9) substance use
- (10) retention

#### Ordinal outcomes

Logistic hierarchical models with a random intercept (assumed normally distributed on the logit scale, with a constant variance) to account for repeated measures within the same participants, a random intercept (assumed normally distributed on the logit scale, with a constant variance) for site, fixed effects for the outcome at baseline, cannabis use, time (12-weeks and 3 month follow up), treatment group, and an interaction term for study time and treatment group. A cumulative logit link will be used to obtain the proportional odds. In the event that proportional odds assumptions are not appropriate, either multinomial logistic regression models or logistic regression models with a logit link will be used with appropriate cut point for outcome categorisations. Treatment effects for each follow-up timepoint will be reported as estimated odds ratio between the two treatment groups with 95% credible intervals and a bayes factor.

- (5) categorical average daily consumption of cigarettes.
- (6) Cravings, strength and frequency
- (6) Minnesota Nicotine Withdrawal scale

## Missing data

Hierarchical Bayesian modelling will be used to perform analyses under an ITT framework assuming missing at random (MAR). All missing outcome and baseline data will be multiply imputed using predictive mean matching and 20 burn-ins, with the number of imputed datasets equal to the percentage of the missing primary outcome responses or 20, whichever is higher. Site, cannabis use, treatment group, variables that are predictive of the primary outcome, and variables that are predictive of drop-out will be used as predictors of the missing data. Initial imputed values will be checked against scoring mechanisms for each questionnaire, and values adjusted as needed prior to modelling. The posterior draws of each model will be combined.

For each outcome, as a sensitivity analysis, if assumptions for MAR are not supported in literature, multiple imputation will be used to impute missing outcome data under varying MNAR assumptions (as determined to be clinically probable).

### Secondary population

The secondary population in the trial is the compliers. The complier average causal effect (CACE) for VPN’s will be used to explore the impact of compliance on the primary outcome. The level of compliance will be defined from retention (secondary outcome 10). The same model to assess the primary outcome will be used, with one difference: *X_ijk_* = 0 (NRT group) or *R*, where *R* is defined as the proportion of participants in the VNP group that adhered to treatment. The value of y is the level of compliance observed, and y is then estimated as if all participants were adherent by scaling y by 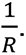.

### Sensitivity analyses

Sensitivity analyses will be used to examine the robustness of results.

1. The impact of the choice of prior on the primary outcome will be examined by running the primary analysis with informative (power) priors with three different weights (0.25, 0.5, 1). The power prior will be based on previous literature [4, 5] that compared VNP and NRT.
2. The primary model will be run again with only complete cases to determine the robustness of the multiple imputation.
3. The primary model will be run with varying MNAR assumptions deemed to be clinically probable.

The sensitivity analysis results will be presented alongside the primary analysis in the format of Table A1 (See Appendix).

### Adverse events

All adverse events will be recorded and reported via descriptive statistics.

### Statistical software

Analyses will be performed using SAS v9.4 (SAS Institute, Cary, North Carolina, USA) or R (R Core Team, 2022) or Stan (Stan Development Team, 2024).

## Data Availability

All data produced in the present study are available upon reasonable request to the authors

## APPENDIX

**Table A1.**
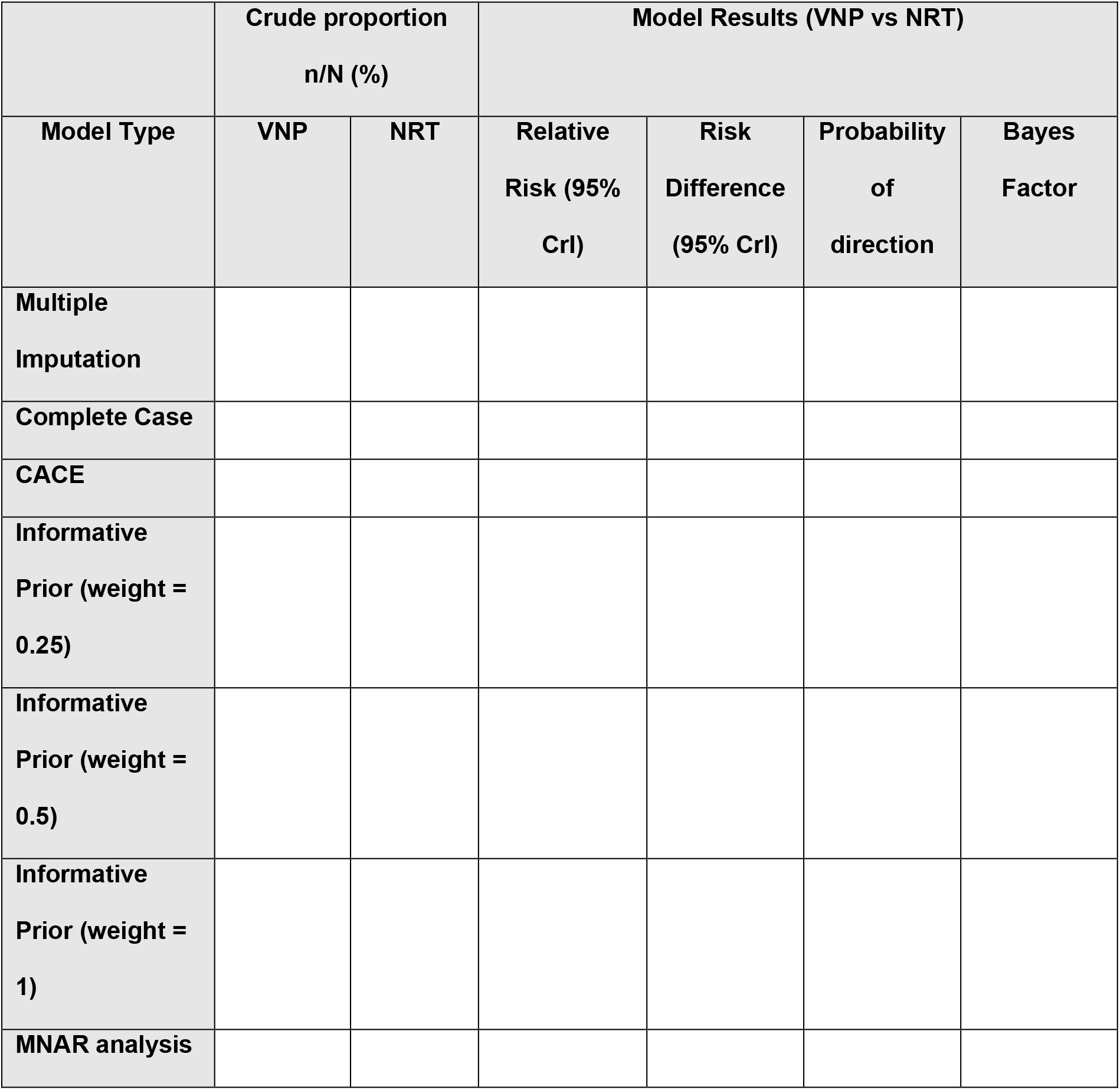
Self-reported 7-day abstinence from smoking.

## Notes

### Competing Interest Statement

The authors have declared no competing interest.

### Funding Statement

The study was funded by New South Wales Health Translational Research Grant Scheme

